# Metabolomic Profiles of Scleroderma-PAH are different than idiopathic PAH and associated with worse clinical outcomes

**DOI:** 10.1101/2021.07.10.21259355

**Authors:** Mona Alotaibi, Junzhe Shao, Michael W. Pauciulo, William C. Nichols, Anna R. Hemnes, Atul Malhotra, Nick H. Kim, Jason X.-J. Yuan, Timothy Fernandes, Kim M. Kerr, Laith Alshawabkeh, Ankit A. Desai, Jeramie D. Watrous, Susan Cheng, Tao Long, Stephen Y. Chan, Mohit Jain

## Abstract

The molecular signature in patients with systemic sclerosis (SSc)-associated pulmonary arterial hypertension (SSc-PAH) relative to idiopathic pulmonary arterial hypertension (IPAH) remain unclear. We hypothesize that patients with SSc-PAH exhibit unfavorable bioactive metabolite derangements compared to IPAH that contribute to their poor prognosis and limited response to therapy. We sought to determine whether circulating bioactive metabolites are differentially altered in SSc-PAH versus IPAH.

Plasma biosamples from 415 patients with SSc-PAH (cases) and 1115 patients with IPAH (controls) were included in the study. Over 700 bioactive metabolites were assayed in plasma samples from independent discovery and validation cohorts using liquid chromatography - mass spectrometry (LC-MS) based approaches. Regression analyses were used to identify metabolites which exhibited differential levels between SSc-PAH and IPAH and associated with disease severity.

From among hundreds of circulating bioactive molecules, twelve metabolites were found to distinguish between SSc-PAH and IPAH, as well as associate with PAH disease severity. SSc-PAH patients had increased levels of fatty acid metabolites including lignoceric acid and nervonic acid, as well as kynurenine, polyamines, eicosanoids/oxylipins and sex hormone metabolites relative to IPAH. In conclusion, SSc-PAH patients are characterized by an unfavorable bioactive metabolic profile that may explain the poor and limited response to therapy. These data provide important metabolic insights into the pathogenesis of SSc-PAH.

**Take Home Message:** Among patients with PAH, those with SSc-PAH suffer disproportionately worse outcomes and disease course. This study represents the most comprehensive analysis of bioactive metabolites profiling comparing two subgroups of PAH. The findings shed light on key differences between SSc-PAH and IPAH that provide important metabolic insight into the disease pathogenesis.

## Introduction

Pulmonary arterial hypertension (PAH) is a debilitating disease of the pulmonary circulation leading to elevated pulmonary arterial pressures and pulmonary vascular resistance. The most common subgroups of PAH are idiopathic pulmonary arterial hypertension (IPAH) and systemic sclerosis associated pulmonary arterial hypertension (SSc-PAH).^1^ Systemic Sclerosis (SSc) is a complex, immunological disease, characterized by autoimmunity, fibrosis of the skin and internal organs, and small vessel vasculopathy. ^2^ PAH is a leading cause of death in patients with SSc with mortality three-fold higher than patients with IPAH. ^3-6^ Despite the comparable end pathology in SSc-PAH and IPAH, SSc-PAH patients have an impaired response to traditional PAH-targeted therapies and carry a worse prognosis relative to other subgroups of PAH, although they may present with milder hemodynamic impairment.^3; 7^ Proposed factors explaining these striking disparities include more pronounced inflammation^8^, autoimmunity^8^, the nature of the underlying vasculopathy^9^ and the ability of the right ventricle to adapt to the increased afterload.^10^ In contrast to IPAH, patients with SSc-PAH have depressed sarcomere function portending worse RV contractility.^11^ However, little is known about the molecular mechanisms underlying these differences. A clearer understanding of the molecular mechanisms underlying SSc-PAH is critical toward better understanding of the disease pathogenesis, including development of prognostic biomarkers and targeted therapies.

Prior mass-spectrometry methods have identified bioactive molecules in circulation.^12; 13^ These molecules include both endogenous compounds (e.g., amino acids, short peptides, nucleic acids, fatty acids, lipids, amines, carbohydrates) and exogenous chemicals that are not naturally produced in the body. Their levels provide integrative information on biological functions and define the phenotypes of biological systems in response to genetic or environmental changes. To date, the study of these bioactive metabolites in PAH have revealed changes in key energetic pathways, including abnormal oxidation products as well as elevated levels of circulating acylcarnitine, glutamate, and TCA cycle intermediates.^14; 15^ Despite these early studies, a deeper understanding of the metabolic alterations between subgroups of PAH and more specifically SSc-PAH has been limited to date.

In this study, we hypothesize that patients with SSc-PAH exhibit unfavorable metabolic derangements which are associated with worse clinical outcomes (i.e low 6MWD, mortality, etc.) compared to IPAH that could explain the rapid decline and disease pathogenesis. We compared hundreds of circulating bioactive metabolites between SSc-PAH and IPAH in independent studies and identified selective derangements between these subgroups that also associate with hemodynamic measures.

## Methods

### Cohorts and sample collection

Plasma samples were obtained from patients with PAH enrolled as part of the National Biological Sample and Data Repository for Pulmonary Arterial Hypertension (PAH Biobank) between October 2012 and December 2017 from 37 US centers (www.pahbiobank.org). The PAH biobank samples were divided *a priori* into discovery and validation cohort (1st validation cohort) based on center from which samples were collected. In the discovery cohort, 310 SSc-PAH patients (cases) and 869 IPAH patients (controls) were included. In the validation cohort (1st validation cohort), 90 SSc-PAH and 213 IPAH patients were included. A second validation cohort (2^nd^ validation cohort) of 15 SSc-PAH patients and 90 IPAH patients was obtained from Vanderbilt Medical Center. Inclusion criteria were confirmed precapillary pulmonary hypertension by right heart catheterization (RHC) (mPAP≥25 mmHg, PCWP≤15 mmHg, PVR >3WU). Diagnosis of SSc-PAH was established clinically based on published criteria.^16^

Plasma samples were obtained from the antecubital fossa and collected in EDTA vacutainer tubes, immediately put-on ice, centrifuged, and stored at -80°C. World Health Organization functional class (WHO FC), 6-minute walk distance (6MWD) and clinical and hemodynamics data were recorded for all patients, using established criteria. All subjects provided informed consent and local research ethics committees approved the study.

### Metabolite Assay

Bioactive metabolites analysis was performed on plasma samples by liquid chromatography - mass spectrometry (LC-MS), using a Vanquish UPLC coupled to high resolution, QExactive orbitrap mass spectrometer (Thermo), similar to as previously described^12; 17^ (details can be found in the online data supplement). Polar metabolites including sugars and organic acids, were assayed using Zic-pHILIC 2.1×150mm 5µm column, and small polar, bioactive lipids were measured using a Phenomenex Kinetex C18 column.^17-26^ Metabolites identified as xenobiotics or detected in <20% of samples were excluded from the analysis, leaving over 700 well-quantified biological metabolites. Qc/Qa analysis was performed as described in data supplement, and spectral data were extracted as previously described.^18-20; 22^ Data were subsequently normalized using batch median normalization metric with correction for median absolute deviation. Following normalization, metabolite peaks were further compressed for multiple adducts and in source fragments. Normalized, aligned, filtered datasets were subsequently used for statistical analyses, as described below.

### Statistical Analysis

Initial group comparisons between SSc-PAH and IPAH patients were performed using t-test or the Mann-Whitney test for continuous variables and the χ^2^ test for categorical variables. Prior to all analyses, metabolite values were natural logarithmically transformed, as needed, and later standardized with mean=0 and SD=1 to facilitate comparisons. Logistic regression analysis was used to determine metabolites that were significantly different between SSc-PAH and IPAH (analysis I). Linear regression was preformed between the significant metabolites from analysis I and 6MWD, FC, mRAP, and PVR in both IPAH and SSc-PAH (analysis II). Sensitivity analysis was performed in IPAH only and SSc-PAH only. Sub-analysis adjusting for immunosuppression medication and steroid use was performed in the significant metabolites. All analyses were performed in models adjusting for age, gender, and body mass index (BMI). To determine significance, a Bonferroni corrected *P* value threshold of 0.05 divided by a conservative estimate of the total number of unique small molecules (i.e., *p*<10^−4^) was used. Lasso regularized regression model was used to build a prediction model and metabolite risk score (MRS) to select the minimum number of metabolites that distinguish between SSc-PAH and IPAH. Statistical analysis was performed with R with RStudio and associated packages.^27^

## Results

### Analysis of Study Cohorts

Baseline demographic, clinical and hemodynamic characteristics and medications for patients enrolled in the study are summarized in **Table 1 and Table e1**. Patients with SSc-PAH were significantly older with female predominance compared to IPAH. At the time of enrollment, SSc-PAH patients had significantly lower mRAP and PVR than IPAH counterparts. Most patients with SSc-PAH were on anti-inflammatory and immunosuppressant therapies at the time of enrollment.

**Table 1.** Patients characteristics. Demographics and clinical features of patients with IPAH and SSc-PAH. Mean±SD, counts or percentages are shown. BMI indicates body mass index; SMWD, six-minute walk distance; mRAP, mean right atrial pressure; PVR, pulmonary vascular resistance.

|  | Discovery cohort |  |  | 1<sup>st</sup> Validation cohort |  |  | 2<sup>nd</sup> Validation cohort |  |  |
| --- | --- | --- | --- | --- | --- | --- | --- | --- | --- |
|  | IPAH | SSc-PAH | *P* | IPAH | SSc-PAH | *P* | IPAH | SSc-PAH | *P* |
| N | 864 | 310 |  | 213 | 91 |  | 32 | 15 |  |
| Female (%) | 663 (76.7) | 267 (86.1) | 0.001 | 166 (77.9) | 82 (90.1) | 0.019 | 24 (75.0) | 14 (93.3) | 0.275 |
| Age (mean (SD)) | 51.90 (18.47) | 64.10 (11.03) | <0.001 | 53.22 (14.95) | 63.74 (10.29) | <0.001 | 42.09 (17.92) | 57.95 (12.72) | 0.004 |
| BMI (mean (SD)) | 30.36 (19.14) | 28.24 (11.40) | 0.068 | 30.78 (9.23) | 27.32 (8.17) | 0.002 | 30.97 (8.83) | 27.08 (4.47) | 0.175 |
| Renal Insufficiency (%) | 32 (3.7) | 28 (9.0) | <0.001 | 11 (5.2) | 7 (7.7) | 0.555 | NA | NA |  |
| Cirrhosis (%) | 13 (1.5) | 5 (1.6) | 1 | 2 (0.9) | 4 (4.4) | 0.125 | NA | NA |  |
| Functional Class (%) |  |  | 0.093 |  |  | 0.51 |  |  | 0.46 |
| I | 46 (7.1) | 14 (5.7) |  | 5 (3.9) | 0 (0.0) |  | NA | NA |  |
| II | 182 (28.3) | 81 (33.1) |  | 42 (32.6) | 20 (36.4) |  | 5 (16.7) | 3 (21.4) |  |
| III | 345 (53.6) | 135 (55.1) |  | 72 (55.8) | 31 (56.4) |  | 22 (73.3) | 11 (78.6) |  |
| IV | 71 (11.0) | 15 (6.1) |  | 10 (7.8) | 4 (7.3) |  | 3 (10.0) | 0 (0.0) |  |
| SMWD (mean (SD)) | 353.91 (138.36) | 312.44 (118.06) | 0.001 | 348.80 (125.44) | 312.30 (130.54) | 0.121 | 293.31 (129.27) | 304.42 (78.73) | 0.79 |
| mRAP (mean (SD)) | 9.16 (5.85) | 8.33 (5.06) | 0.028 | 8.38 (4.99) | 7.48 (4.94) | 0.154 | 8.57 (6.06) | 6.93 (4.57) | 0.375 |
| PVR (mean (SD)) | 10.74 (6.84) | 8.50 (5.13) | <0.001 | 12.18 (6.38) | 8.48 (4.06) | <0.001 | 55.76 (213.47) | 9.06 (4.62) | 0.405 |
| Prostanoids use (%) | 415 (48.0) | 130 (41.9) | 0.075 | 83 (39.0) | 25 (27.5) | 0.074 | 12 (37.5) | 4 (26.7) | 0.689 |

### Metabolites distinguishing between SSc-PAH and IPAH

Circulating levels of 94 bioactive metabolites across broad chemical classes, including bioactive lipids and polar metabolites, distinguished SSc-PAH from IPAH in both discovery and 1^st^ validation cohorts at a ‘metabolome wide’ statistical threshold of *p*<10^−4^ after correction for confounders including age, gender and BMI (**Figure 1**). These metabolites included alterations in fatty acid oxidation and eicosanoids metabolism, steroid hormones, kynurenine pathway, polyamine and pyrimidine pathways (**Figure 1**). Orthogonal partial least squares-discriminant analysis (OPLS-DA) plots showed clear separation between SSc-PAH and IPAH (**Figure 1e**).

**Figure 1:**
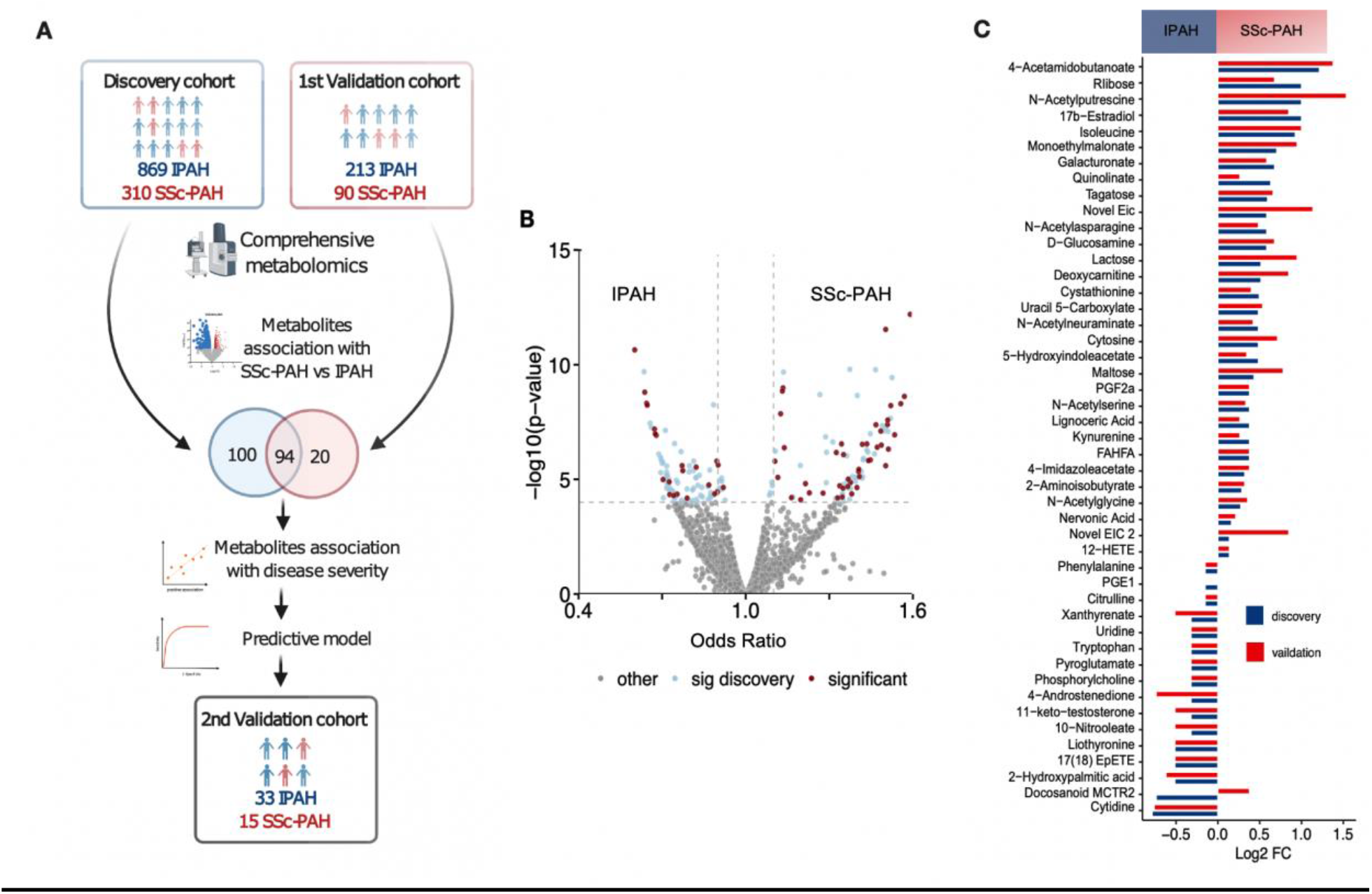
Metabolites distinguishing between SSc-PAH and IPAH. **A**. Study Flow chart Summary of study workflow and data interpretation. **B**. Volcano plot of metabolites distinguishing SSc-PAH from IPAH in the discovery and validation cohorts **D**. Average metabolite levels in SSc-PAH and IPAH patients for 47 metabolites found to significantly distinguish patients with SSc-PAH from IPAH. Values plotted are log2 fold change. Negative values indicate metabolites at lower levels in patients with SSc-PAH and positive values indicate metabolites at higher levels in patients with SSc-PAH. FC indicate fold change.

### Determining Metabolites which most differentiate SSc-PAH from IPAH

To identify a minimal set of metabolites able to distinguish between SSc-PAH and IPAH, we performed regularized regression analysis. A metabolite risk score (MRS) comprised of 12 metabolites was able to distinguish IPAH from SSc-PAH with discrimination at an AUC of 0.86 (95% CI: 0.82-0.91). This MRS was validated in an independent validation cohort (2^nd^ validation cohort) of 33 IPAH patients and 15 SSc-PAH patients from Vanderbilt Medical Center with an AUC of 0.76 (95% CI: 0.60-0.92) (**Figure 2**). These 12 metabolites were associated with markers of disease severity. 11 of the selected metabolites remained significant after adjusting for immunosuppression medication and steroid use.

**Figure 2:**
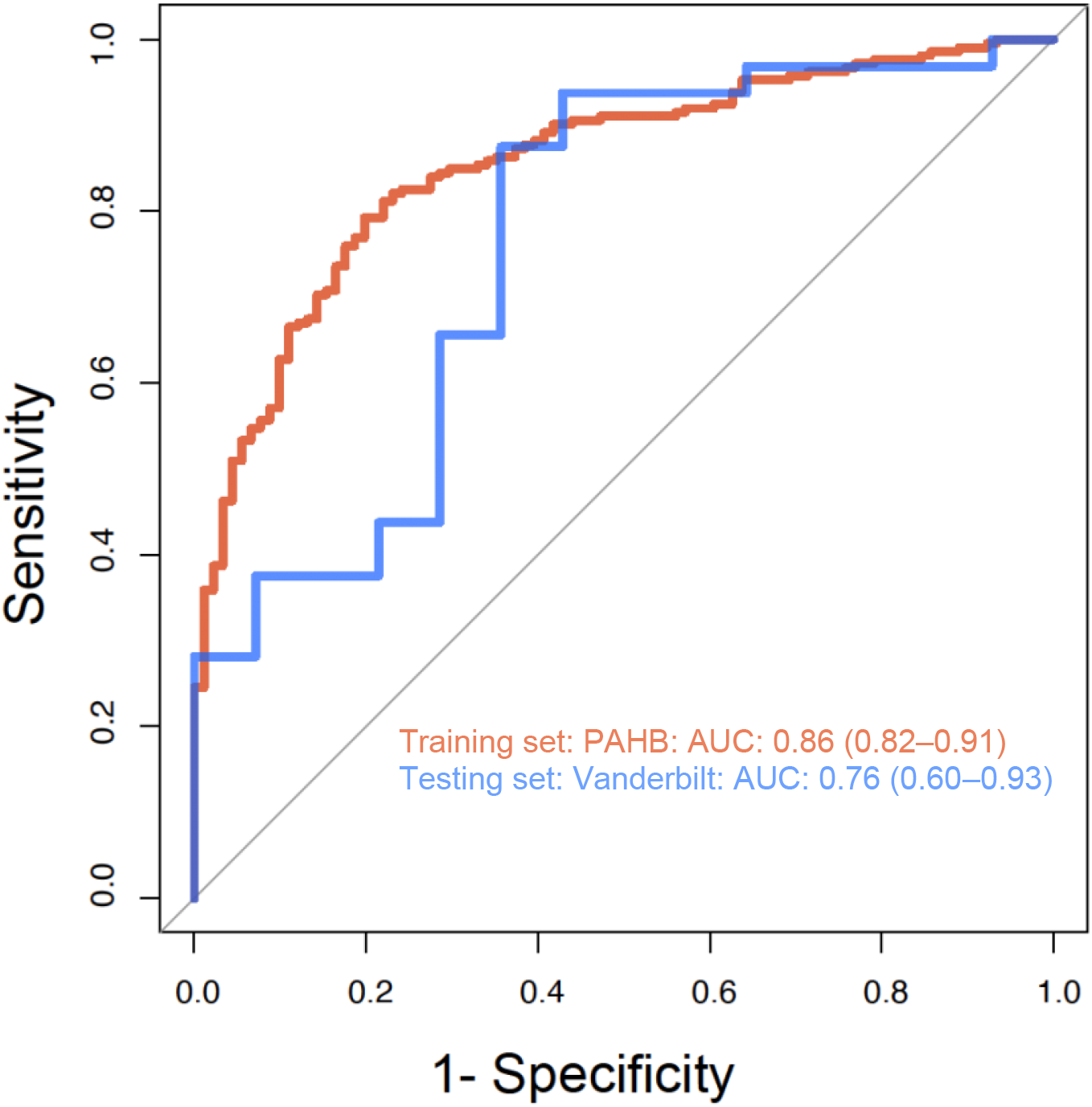
ROC curve. Receiver-operating characteristic curves showing the performance of the model in distinguishing IPAH from SSc-PAH using 12 metabolites. Blue curve represent the discovery cohort and the red curve represent the independent validation cohort (2^nd^ validation cohort). AUC indicate area under the curve, and CI, confidence interval.

### Metabolites differentiating SSc-PAH from IPAH and associated with disease severity

To determine if distinguishing metabolites may contribute to worsening disease prognosis, we next performed association of the 94 metabolite biomarkers with available clinical markers of disease severity and hemodynamic parameters, including 6MWD, FC, mRAP, and PVR. As shown in **Table 2** and **Figure 3**, 30 metabolites were significantly associated with at least one measurement of disease severity (*p*<0.05 for each metabolite). This includes fatty acid metabolites such as very long chain saturated fatty acids (VLCSFA) and monounsaturated unsaturated fatty acids (MUFA), several pro-inflammatory eicosanoids, kynurenine, polyamines and sex hormones. Additionally, novel eicosanoids and fatty acid ester of hydroxyl fatty acid (FAHFA) emerged as markers of disease severity and were significantly higher in SSc-PAH.

**Table 2:** Metabolites distinguishing SSc-PAH from IPAH and associated with markers of disease severity

| Metabolite | Metabolic Pathway | Discovery |  |  | Validation |  |  |
| --- | --- | --- | --- | --- | --- | --- | --- |
|  |  | *P* | FC | OR | *P* | FC | OR |
| 2-deoxy-d-glucose/ d-glucosamine | Amino sugar and nucleotide sugar metabolism | 7.00E-07 | 1.5 | 1.33 | 2.00E-02 | 1.6 | 1.31 |
| N-Acetylneuraminate | Amino sugar and nucleotide sugar metabolism | 2.00E-09 | 1.4 | 1.57 | 1.00E-03 | 1.34 | 1.66 |
| 17(18) EpETE | Arachidonic acid metabolism | 4.00E-05 | 0.7 | 0.89 | 1.00E-02 | 2.5 | 0.88 |
| Novel Eic | Arachidonic acid metabolism | 1.00E-08 | 1.5 | 1.13 | 3.00E-03 | 2.2 | 1.13 |
| Novel Eic 2 | Arachidonic acid metabolism | 1.00E-06 | 1.1 | 1.35 | 1.00E-03 | 1.8 | 1.55 |
| PGE1 | Arachidonic acid metabolism | 2.00E-05 | 0.9 | 0.8 | 7.00E-03 | 1 | 0.84 |
| PGF2a | Arachidonic acid metabolism | 4.00E-07 | 1.3 | 1.14 | 2.00E-02 | 1.3 | 1.11 |
| N-Acetylputrescine | Arginine and proline metabolism | 1.00E-12 | 2 | 1.69 | 3.00E-04 | 2.9 | 1.7 |
| 4-Acetamidobutanoate | Arginine and proline metabolism | 2.00E-14 | 2.32 | 1.72 | 3.00E-05 | 1.86 | 2.6 |
| Acetylserine | Cysteine and methionine metabolism | 5.00E-06 | 1.3 | 1.4 | 8.00E-03 | 1.26 | 1.51 |
| Cystathionine | Cysteine and methionine metabolism | 4.00E-05 | 1.41 | 1.28 | 3.00E-02 | 1.32 | 1.32 |
| FAHFA | Fatty Acid metabolism | 8.00E-08 | 1.3 | 1.5 | 8.00E-03 | 1.3 | 1.54 |
| Lignoceric Acid | Fatty Acid metabolism | 1.00E-07 | 1.3 | 1.5 | 3.00E-03 | 1.2 | 1.57 |
| Nervonic Acid | Fatty Acid metabolism | 4.00E-07 | 1.12 | 1.5 | 2.00E-03 | 1.16 | 1.58 |
| Tagatose | Galactose metabolism | 4.00E-05 | 1.51 | 1.38 | 3.00E-03 | 1.58 | 1.63 |
| 4-Imidazoleacetate | Histidine metabolism | 6.00E-05 | 1.25 | 1.35 | 3.00E-03 | 1.3 | 1.58 |
| 2-Aminoisobutyrate | Leucine, Isoleucine and Valine Metabolism | 1.00E-05 | 1.22 | 1.37 | 3.00E-02 | 1.25 | 1.38 |
| Cytosine | Pyrimidine metabolism | 8.00E-05 | 1.4 | 1.2 | 1.00E-03 | 1.64 | 1.32 |
| Uracil 5-carboxylate | Pyrimidine metabolism | 6.00E-12 | 1.4 | 1.62 | 4.00E-04 | 1.45 | 1.62 |
| 17α-testosterone | Steroid hormones metabolism | 1.00E-05 | 0.8 | 0.87 | 1.00E-03 | 0.7 | 0.75 |
| 17b-Estradiol | Steroid hormones metabolism | 1.0E-09 | 2 | 1.13 | 3.00E-03 | 1.8 | 1.12 |
| Kynurenine | Tryptophan metabolism | 2.00E-13 | 1.3 | 1.7 | 2.00E-03 | 1.2 | 1.57 |
| Quinolate | Tryptophan metabolism | 1.00E-05 | 1.55 | 1.38 | 2.00E-02 | 1.2 | 1.41 |
| Tryptophan | Tryptophan metabolism | 1.00E-09 | 0.8 | 0.68 | 4.00E-03 | 0.8 | 0.66 |
| Xanthurenate | Tryptophan metabolism | 3.00E-06 | 0.8 | 0.77 | 2.00E-02 | 0.7 | 0.78 |
| 5-Hydroxyindoleacetate | Tryptophan metabolism | 2.00E-13 | 1.7 | 1.34 | 6.00E-03 | 1.27 | 1.4 |
| N-Acetylasparagine | Alanine, aspartate and glutamate metabolism | 7.00E-06 | 1.5 | 1.4 | 9.00E-03 | 1.4 | 1.4 |
| Deoxycarnitine | Fatty Acid metabolism | 2.00E-05 | 1.43 | 1.3 | 1.00E-02 | 1.8 | 1.4 |
| Monoethylmalonate | Fatty Acid metabolism | 4.00E-08 | 1.63 | 1.4 | 2.00E-05 | 1.93 | 2 |
| Nitrooleate | Fatty Acid metabolism | 2.00E-05 | 0.6 | 0.9 | 2.00E-02 | 0.7 | 0.9 |

**Figure 3:**
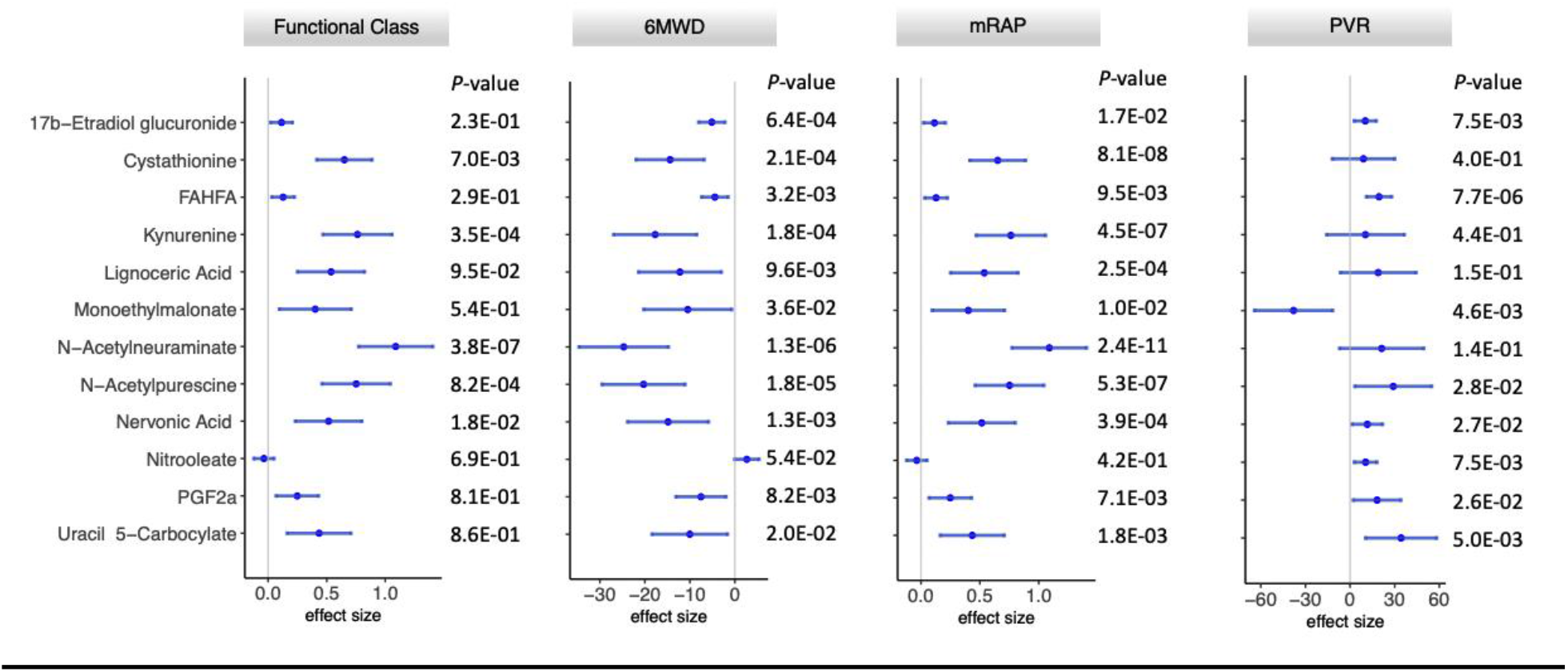
Forrest Plot for the 12-MRS metabolites and association with clinical variables.

## Discussion

In this report, we provide novel evidence that patients with SSc-PAH have significant bioactive metabolic alterations compared to those with IPAH. We assayed hundreds of circulating bioactive metabolites using LC-MS approaches in 415 SSc-PAH patients and 1115 IPAH controls in independent discovery and validation cohorts. We identified a set of bioactive metabolite biomarkers independently differentiating SSc-PAH from IPAH and associated with disease severity, after adjusting for age, gender, BMI and medications. In combination, these biomarkers were able to distinguish SSc-PAH patients from IPAH with high degree of accuracy. These findings provide molecular insight into the heterogeneity between PAH subgroups and could explain in part the worse prognosis and response to therapy in patients with SSc-PAH.

Our novel findings, which highlight 12 metabolites from among hundreds assayed, are significant both pathophysiologically and clinically. Pathophysiologically, this work sheds light on potential different mechanisms between SSc-PAH and IPAH. Clinically, it suggests that these biomarkers could be explored as potential prognostic tools and therapeutic targets.

We observed novel associations between saturated and unsaturated fatty acids that were significantly higher in patients with SSc-PAH and correlated with worse disease markers. Very-long-chain saturated fatty acids (VLCSFAs) are group of saturated fatty acids with a chain length of ≥20 carbon atoms. They have distinct functions when compared to long chain saturated fatty acids, and are involved in liver homeostasis, retinal function, and anti‐inflammatory functions^30^. Little is known about the role of VLCSFA in relation to pulmonary vascular pathology. In fact, this is the first observation to our knowledge to associate VLCSFA with PAH. In our study, VLCSFAs were positively associated with PVR, mRAP and negatively associated with 6MWD. Monounsaturated fatty acids (MUFA), such as nervonic acid, are involved in many physiological processes, including energy metabolism, antioxidant reactions and apoptosis.^31^ In previous reports, nervonic acid was positively associated with greater congestive heart failure, poor performance and increased risk of cardiovascular mortality.^32; 33^

In our study, patients with SSc-PAH had more significant alterations in several pathways linked to endothelial cells dysfunction and RV dysfunction like kynurenine pathway, polyamines and spermine metabolism, eicosanoids, sex hormones among others, despite having better hemodynamic profile at the time of enrollment. Although some of these markers were described in cardiovascular disease or pulmonary hypertension, this is the first-time showing increase levels in SSc-PAH.

Levels of kynurenine, an immune signaling molecule, correlated with resting pulmonary artery pressure in unexplained dyspnea patients and patients with both mild and severe PH.^15^ Plasma levels of polyamines metabolites including N-acetylputrescine was associated with chronic inflammation and previous reports showed increased levels of acetylputrescine in animal models of PAH.^34^ Interestingly, the administration of polyamines inhibitors in monocrotaline rats prevented the development of pulmonary hypertension and RV dysfunction^35^, suggesting this could have therapeutic implications as well in this subgroup specifically. Several pro-inflammatory eicosanoids such as, 15-HETE, 17(18)-EpETE and prostaglandins have been implicated in the pathogenesis of PAH and associated with disease severity.^36^

Rhodes et al^14^ was among the first to use comprehensive LC-MS based metabolomics platform to identify discriminative and prognostic metabolites in PAH. They identified a set of 20 metabolites discriminative between IPAH and healthy and diseased controls. Levels of these metabolites were similar between SSc-PAH and IPAH in our study. Among the prognostic metabolites in IPAH, levels of acetamidobutanoate and acetylputerscine were associated with mortality in the Rhodes study and were significantly elevated in SSc-PAH in our cohorts relative to IPAH. This supports our hypothesis that these metabolites could contribute to the worse outcomes in SSc-PAH.

A few studies have used circulating metabolites to determine potential metabolic pathways altered in scleroderma (with or without PH).^28; 29^ To date however, these studies have been limited by sample size as well as independent validation and did not compare between subgroups of PAH. A comparison between 8 patients with scleroderma without PAH and 10 patients with scleroderma and PAH using nuclear magnetic resonance (NMR) techniques identified increase in glycolysis and altered fatty acid profiles in patients with scleroderma and PAH.^29^ None of our top differentiating metabolites were measured in this study, mainly related to technical differences (use of NMR vs LC-MS) and small sample size. Thus, a strength of our work is the application of broad plasma bioactive metabolites analysis in large independent cohorts of IPAH and SSc-PAH. We understand that it is important to compare between scleroderma-PAH and scleroderma without PAH to ascertain that these differences are not due to scleroderma only. However, this is beyond the scope of this work, and we hope to address this in the future. We believe that choosing metabolites that associate with hemodynamic measures and disease severity is suggestive of PH related biomarkers.

Even though scleroderma can be easily distinguishable in most cases clinically from IPAH, our goal was not to develop a diagnostic tool, rather to identify key metabolites distinguishing these two subtypes that could shed light on the pathogenesis between SSc-PAH and IPAH. Our hope is that in the future, this could be explored more for potential therapeutic targets.

This study has several limitations. Importantly, given the study design, adjustment for all potential confounders between IPAH and SSc-PAH remains difficult. It is possible that medications such as anti-inflammatory and immunosuppressants, demographic features including gender, smoking and other comorbidities may contribute to metabolic changes between IPAH and SSc-PAH. We tried our best adjusting for these factors in the analysis. Finally, while metabolite markers were found to distinguish SSc-PAH from IPAH and independently associate with disease severity, establishing a clear *causal* relationship for the role of these metabolic pathways in SSc-PAH will require independent studies in experimental model systems. Despite these acknowledged limitations, we believe that our findings provide important scientific insight on the metabolic alterations present in SSc-PAH and their potential role in disease pathobiology.

## Conclusion

Our findings suggest that despite the lack of distinctive pathologic features, SSc-PAH is characterized by significant metabolomic alterations compared to IPAH that may contribute to worsening disease and poor response to therapy. Moreover, our study suggests that metabolite levels may distinguish IPAH from SSc-PAH and therefore, different pathways maybe driving the pathogenesis of PAH in these two groups. This observation may lead to much needed novel therapeutic strategies in SSc-PAH patients.

## Data Availability

all data referred to in the manuscript is available in the manuscript and supplementary materials.

## Acknowledgments

Samples and/or Data from the National Biological Sample and Data Repository for PAH, which receives government support under an investigator-initiated grant (R24 HL105333) awarded by the National Heart Lung and Blood Institute (NHLBI) were used in this study. We thank contributors, including the Pulmonary Hypertension Centers who collected samples used in this study, as well as patients and their families, whose help and participation made this work possible.

## Abbreviations list

BMI: Body mass index
EpETE: Epoxyeicosatetraenoic acid
FAHFA: Fatty acid ester of hydroxyl fatty acid
HETE: Hydroxyeicosanoid
IPAH: Idiopathic pulmonary arterial hypertension
LC-MS: Liquid chromatography - mass spectrometry
mRAP: Mean right atrial pressure
MRS: Metabolite risk score
MUFA: Monounsaturated unsaturated fatty acids
PAH: Pulmonary arterial hypertension
PVR: Pulmonary vascular resistance
RHC: Right heart catheterization
SMWD: 6-minute walk distance
SSc: Systemic sclerosis
SSc-PAH: Systemic sclerosis associated pulmonary arterial hypertension
VLCSFA: Very long chain saturated fatty acids
WHO FC: World Health Organization functional class

