## Supplemental data for "Metabolomic Profiles of Scleroderma-PAH are different than idiopathic PAH and associated with worse clinical outcomes"

**Online Data Supplements**

### Bioactive Metabolites Derangements Differentiating Idiopathic and Scleroderma Associated Pulmonary Arterial Hypertension

Mona Alotaibi MD[1], Junzhe Shao[2], Michael W. Pauciulo MBA[3,4], William C. Nichols PhD[3,4], Anna R. Hemnes MD[5], Atul Malhotra MD[1], Nick H. Kim MD[1], Jason X.-J. Yuan MD PhD[1], Timothy Fernandes MD[1], Kim M. Kerr MD[1], Laith Alshawabkeh MD[6], Ankit A. Desai MD[8], Jeramie D. Watrous PhD[7], Susan Cheng MD MPH MMsc[9], Tao Long PhD[7], Stephen Y. Chan MD PhD[10], Mohit Jain MD PhD[7]

**eMethods:**

*Sample preparation*

Plasma samples were thawed at 4C overnight; 20 µL of plasma was transferred into a 96-well extraction plate. Proteins were precipitated with the addition of 80 µL of ice-cold extraction solvent (consisting of ethanol for reverse phase method and a mixture of 45:45:10 methanol:ACN:water for the HILIC method), as described.[1-3] Plates were sealed, vortexed and centrifuged to precipitate proteins with the supernatant containing extract metabolites pipetted into a clean microtiter plate for LC-MS analysis. For the reverse phase methods, samples were passed across a reverse phase SPE column (Phenomenex 8B-S199-UB), and lipid species eluted using 1ml methanol, as described[1,3], before being transferred into a clean microtiter plate for LC-MS analysis.

*Metabolite Assay*

Bioactive metabolites analysis was performed on plasma samples using state of the art LC-MS, using a Vanquish UPLC coupled to high resolution, QExactive orbitrap mass spectrometer (Thermo). Multiple complementary LC methods were performed, each optimized to capture a chemical subset of bioactive metabolites, including polar molecules like sugars and organic acids (using Zic-pHILIC 2.1x150mm 5um column) and small polar, bioactive lipids (using a Phenomenex Kinetex C18 column). All generated spectral data underwent daily Qc/Qa analysis, as described in the section below. Data was extracted using image processing and machine learning based spectral optimization. Each sample MS data file was initially represented as an image using mass to charge and retention time coordinates. A watershed algorithm was applied to image files using peak apex density as a guide to define regions containing putative spectral peaks. An optimized neural network was subsequently applied to all putative spectra peaks to isolate true spectral peaks from ‘false’ background signals. Following data extraction, spectral peaks were cross aligned among datasets using landmark based algorithms to allow for comparison of signals among cohorts. Data was subsequently normalized to account for plate-to-plate variation using a simple batch median normalization metric with correction for median absolute deviation. Following normalization, metabolite peaks were further compressed for multiple adducts and in source fragments. Normalized, aligned, filtered datasets were subsequently used for statistical analyses, as described below. Metabolites were annotated by using an in-house library of commercially available standards or MS/MS fragmentation patterns.

*QC/QA of Spectral data.*

Qc/Qa of data has been standardized using a panel of deuterated internal standards as well as interval pooled plasma samples to monitor fluctuations in extraction efficiency, instrument sensitivity, matrix artifact and mass accuracy. Any samples not meeting Qc/Qa thresholds underwent reinjection. The assay was highly reproducible over several days and independent measures with median coefficient of variance (CV) across analytes at low standard concentrations of 0.15 ng.

Table 1e. Supplements: Medications

| | Discovery cohort | | | 1st Validation cohort | | |
|---|---|---|---|---|---|---|
| | IPAH | SSc-PAH | *P* | IPAH | SSc-PAH | *P* |
| N | 864 | 310 | | 213 | 91 | |
| Warfarin (%) | 326 (37.7) | 41 (13.2) | <0.001 | 93 (43.7) | 18 (19.8) | <0.001 |
| Heparin (%) | 3 (0.3) | 2 (0.6) | 0.855 | 213 (100.0) | 91 (100.0) | NA |
| Corticosteroid (%) | 57 (6.6) | 70 (22.6) | <0.001 | 34 (16.0) | 30 (33.0) | 0.001 |
| Aspirin (%) | 180 (20.8) | 105 (33.9) | <0.001 | 43 (20.2) | 32 (35.2) | 0.009 |
| NSAID (%) | 53 (6.1) | 30 (9.7) | 0.05 | 23 (10.8) | 10 (11.0) | 1 |
| Immunosuppressive medications (%) | 15 (1.7) | 70 (22.6) | <0.001 | 5 (2.3) | 20 (22.0) | <0.001 |
| Synthetic Thyroid (%) | 158 (18.3) | 95 (30.6) | <0.001 | 49 (23.0) | 40 (44.0) | <0.001 |

**Figure 1e. Orthogonal partial least squares-discriminant analysis (OPLS-DA) plots analysis of bioactive lipids and HILIC metabolites.**

A. Scores plot of the OPLS-DA model for bioactive lipids metabolites.
B. Scores plot of the OPLS-DA model for the HILIC positive metabolites.
C. Scores plot of the OPLS-DA model for the HILIC negative metabolites.

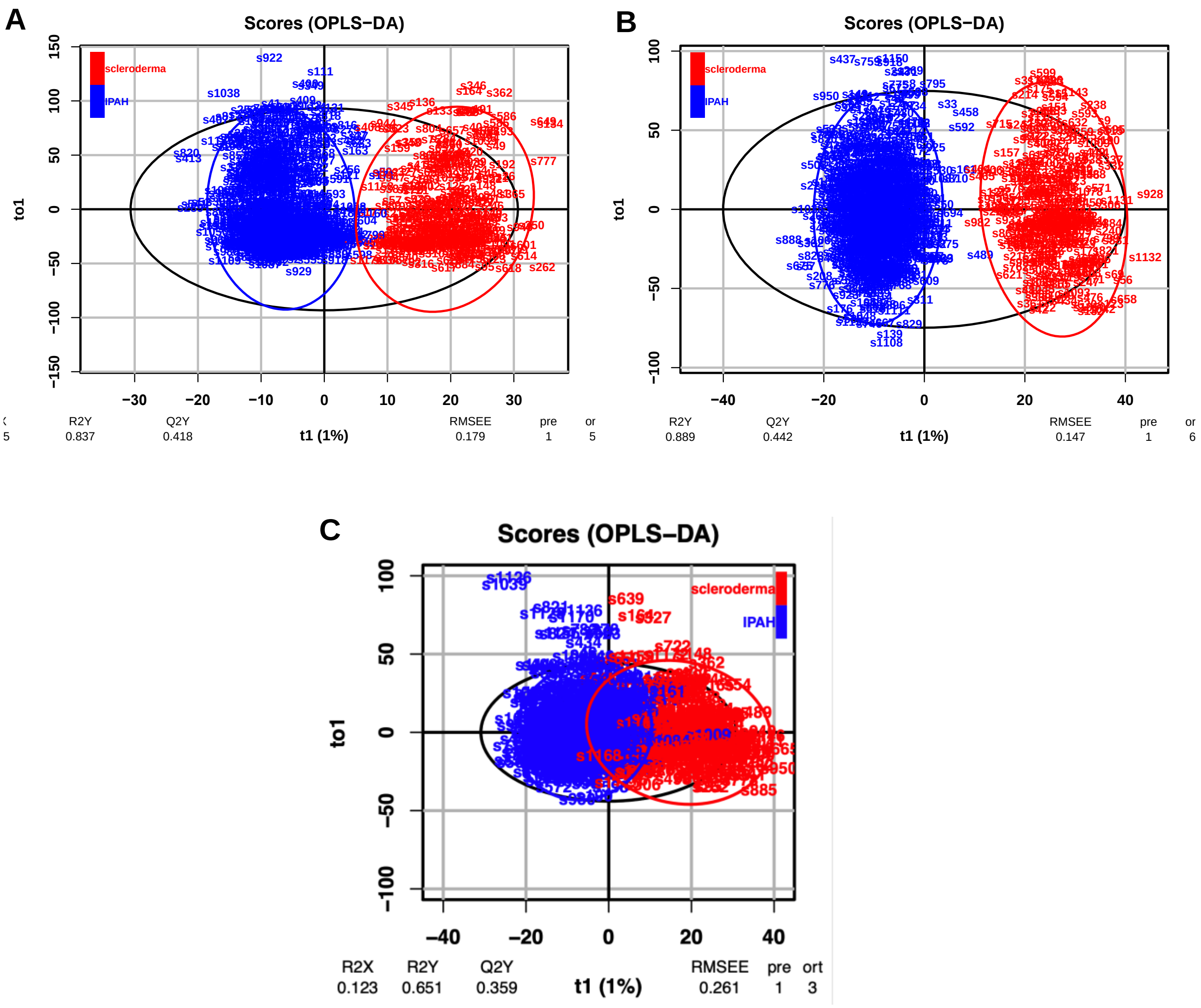
